# Estimated surge in hospitalization and intensive care due to the novel coronavirus pandemic in the Greater Toronto Area, Canada: a mathematical modeling study with application at two local area hospitals

**DOI:** 10.1101/2020.04.20.20073023

**Authors:** Sharmistha Mishra, Linwei Wang, Huiting Ma, Kristy CY Yiu, J. Michael Paterson, Eliane Kim, Michael J Schull, Victoria Pequegnat, Anthea Lee, Lisa Ishiguro, Eric Coomes, Adrienne Chan, Mark Downing, David Landsman, Sharon Straus, Matthew Muller

## Abstract

**Background:** A hospital-level pandemic response involves anticipating local surge in healthcare needs.

**Methods:** We developed a mechanistic transmission model to simulate a range of scenarios of COVID-19 spread in the Greater Toronto Area. We estimated healthcare needs against 2019 daily admissions using healthcare administrative data, and applied outputs to hospital-specific data on catchment, capacity, and baseline non-COVID admissions to estimate potential surge by day 90 at two hospitals (St. Michael’s Hospital [SMH] and St. Joseph’s Health Centre [SJHC]). We examined fast/large, default, and slow/small epidemics, wherein the default scenario (R0 2.4) resembled the early trajectory in the GTA.

**Results:** Without further interventions, even a slow/small epidemic exceeded the city’s daily ICU capacity for patients without COVID-19. In a pessimistic default scenario, for SMH and SJHC to remain below their non-ICU bed capacity, they would need to reduce non-COVID inpatient care by 70% and 58% respectively. SMH would need to create 86 new ICU beds, while SJHC would need to reduce its ICU beds for non-COVID care by 72%. Uncertainty in local epidemiological features was more influential than uncertainty in clinical severity. If physical distancing reduces contacts by 20%, maximizing the diagnostic capacity or syndromic diagnoses at the community-level could avoid a surge at each hospital.

**Interpretation:** As distribution of the city’s surge varies across hospitals over time, efforts are needed to plan and redistribute ICU care to where demand is expected. Hospital-level surge is based on community-level transmission, with community-level strategies key to mitigating each hospital’s surge.

## INTRODUCTION

The COVID-19 pandemic caused by the SARS-Cov2 virus has led to over 1,914,916 detected cases and 123,010 deaths by April 15, 2020 (1). By March 6, 2020, there was direct evidence of local onward transmission in Canada (2). Local transmission refers to acquisition within a geographical locale (in this case, within Canada) but without a direct link to a travel-acquired case. Early evidence from China suggests that among patients diagnosed with COVID-19, 13.8% develop severe disease and 6.1% develop critical illness (3). Thus, an important component of responding to local onward transmission is preparing for a surge in inpatient and intensive care needs for patients with COVID-19 (4-6).

In the Canada’s healthcare system, national, provincial, and local public health agencies provide guidance surrounding pandemic preparedness in the clinical setting, with implementation conducted within each city’s health-care facilities. Indeed, decentralized implementation and hospital-level decision-making has played a major role in the current outbreak (7). Hospital-level pandemic planning teams need to integrate information on their local bed capacity, baseline admissions, and anticipated surge to help prepare their respective hospitals, including workforce planning, within the city, regional, and provincial-level responses (5).

To support hospital-level pandemic planning in the Greater Toronto Area (GTA), we developed an epidemic model and used publicly available data and provincial administrative healthcare data to simulate the range of plausible epidemic trajectories and hospital care needs that may be anticipated for the GTA. We then applied outputs from the epidemic model to hospital-specific data to estimate the early trajectory and daily volume of inpatient and intensive care surge at two downtown, acute-care hospitals in the GTA.

## METHODS

### Study Setting

The GTA has a population of 6 million and includes five regions (8-11) with 40 acute care hospitals (12). By March 20, 2020 there were 266 diagnosed cases of COVID-19 in the GTA (13-18). St. Michael’s Hospital (quaternary care) and St. Joseph’s Health Centre (tertiary care) are part of Unity Health Toronto, a network of two acute care and one long-term continuing care facility. The Unity Health Toronto COVID-19 Incident Management Team was formed on January 27, 2020 and requested rapid modeling to estimate potential surge in health-care needs at each hospital.

### Model design

We developed a deterministic, compartmental, mathematical model of SARS-Cov-2 person-to-person transmission, and simulated a closed population (no births or deaths) over a 300-day period. For the current analyses, we did not stratify the modeled population by age and thus, we assumed a homogenous population. **Figure 1** depicts the model structure, where the biological component follows a susceptible-exposed-infectious-recovered system, and the health-care component includes admissions through inpatient and intensive care units. The model was written in R scripting language (source code available at our GitHub Repository (19)) and is detailed in **Appendix 1**. A R shiny user-interface was created for the model (20).

**Figure 1.**
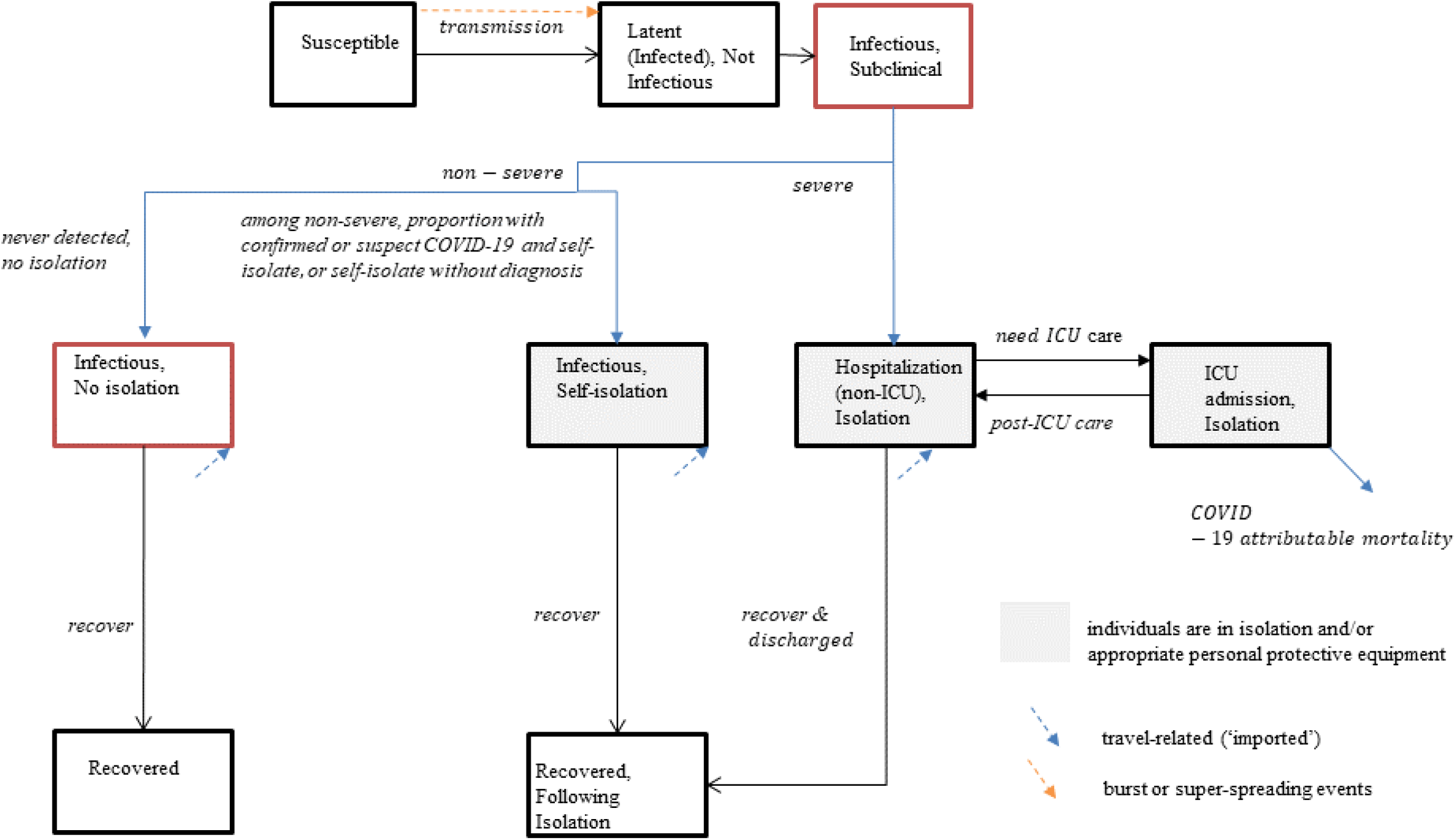
Transmission model structure. Compartments represent health-states, with transitions between health-states in a stable population of fixed size. A proportion of individuals infected with SARS-Cov-2 develop severe COVID-19 and require hospitalization. Among individuals with non-severe COVID-19, a proportion self-isolate after receiving a diagnosis of confirmed or syndromic COVID-19 or may self-isolate without a diagnosis; the remainder do not self-isolate. Only a subset of individuals with non-severe COVID-19 receive a confirmed diagnosis if they undergo testing. Individuals in the infectious health-states may pass the virus on to others. We assume that individuals in self-isolation or hospital-isolation cannot pass on the virus, but superspreading events are included to capture community, long-term care, and nosocomial (hospital-acquired) clusters of transmission events. Abbreviation: ICU: intensive care unit.

Parameter values and their data sources are shown in **Table 1. Appendix 1** details the biological, epidemiological, and clinical severity parameters; internal validity checks (case fatality proportions and serial intervals); and epidemic constraints.

**Table 1.**
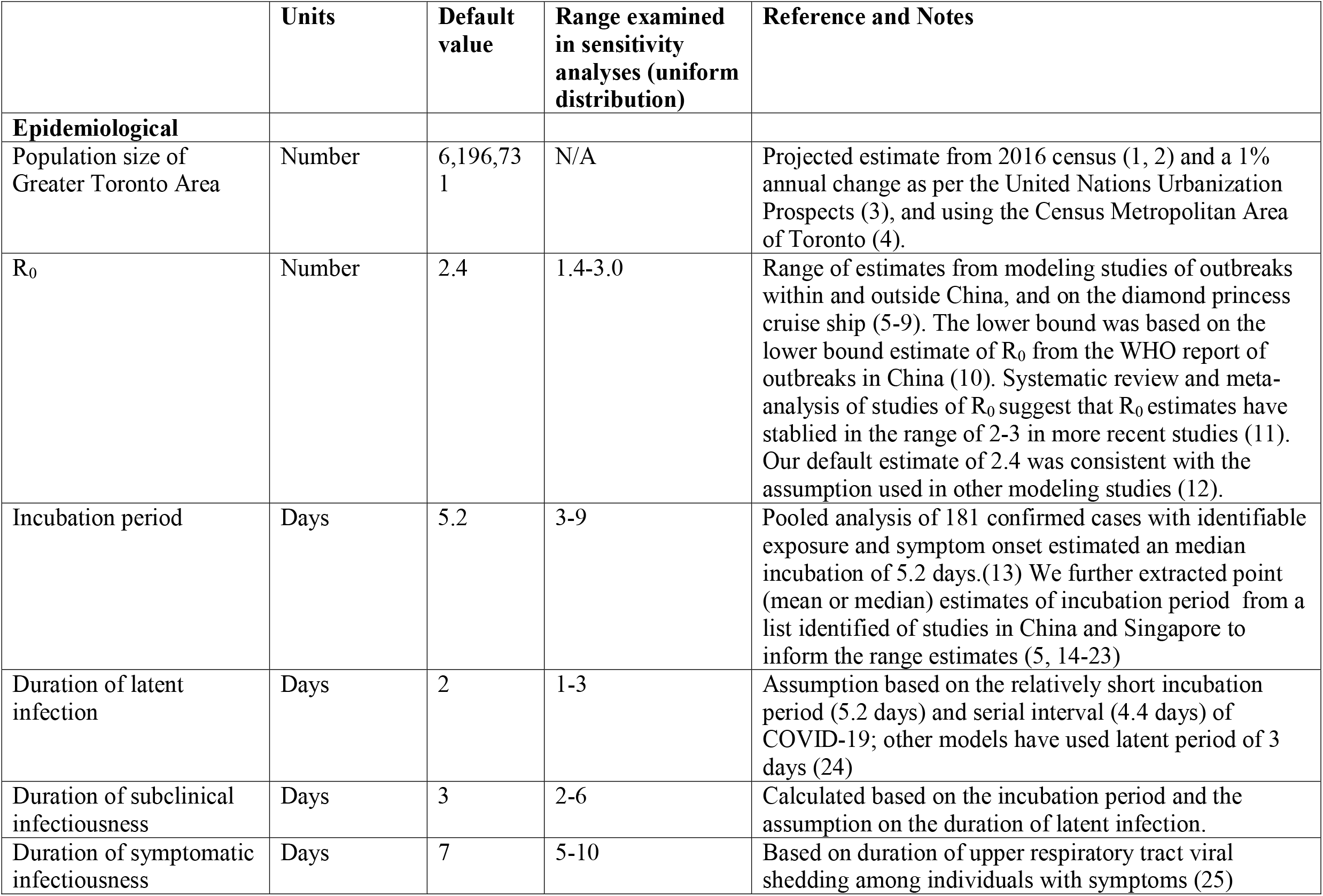
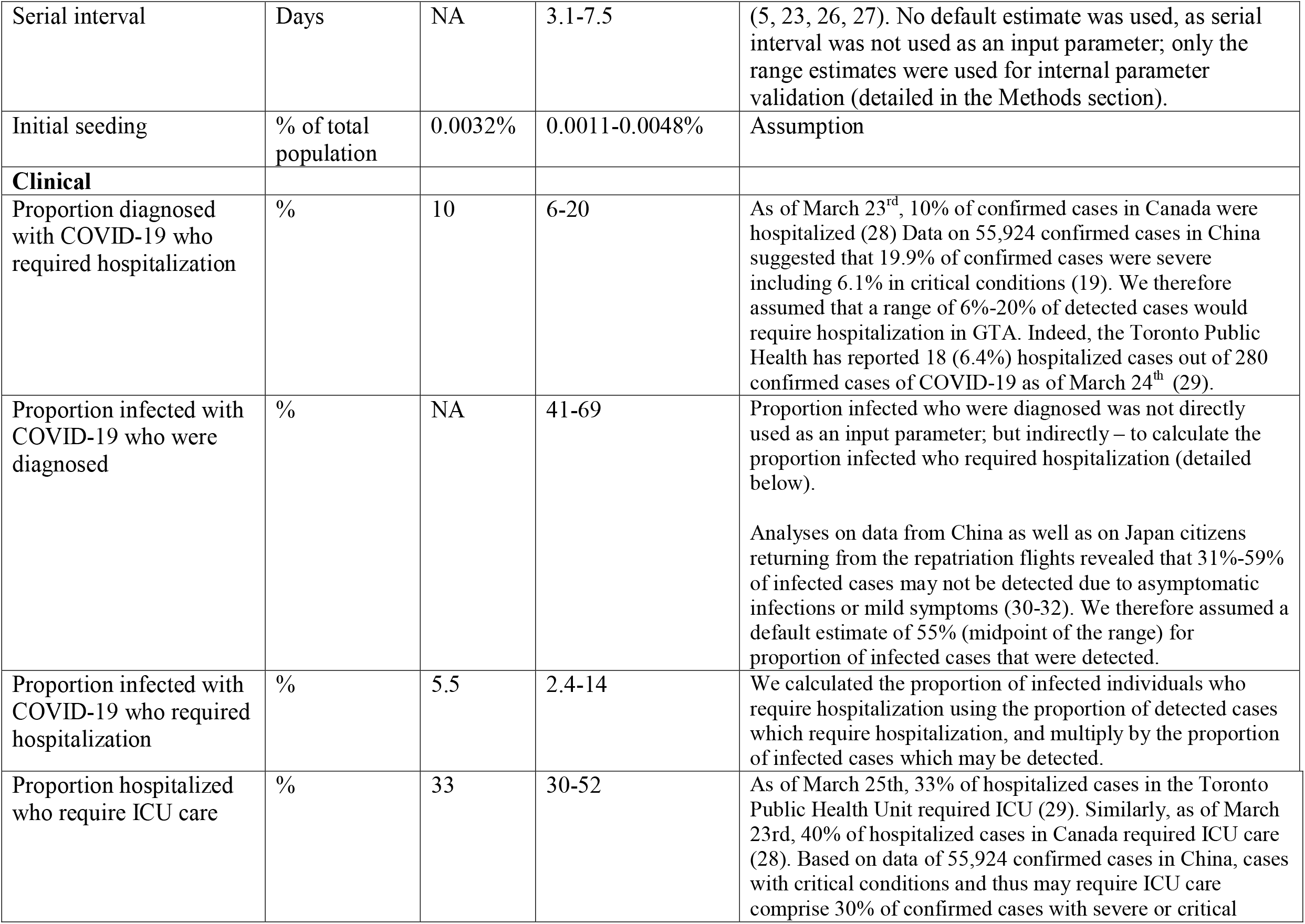
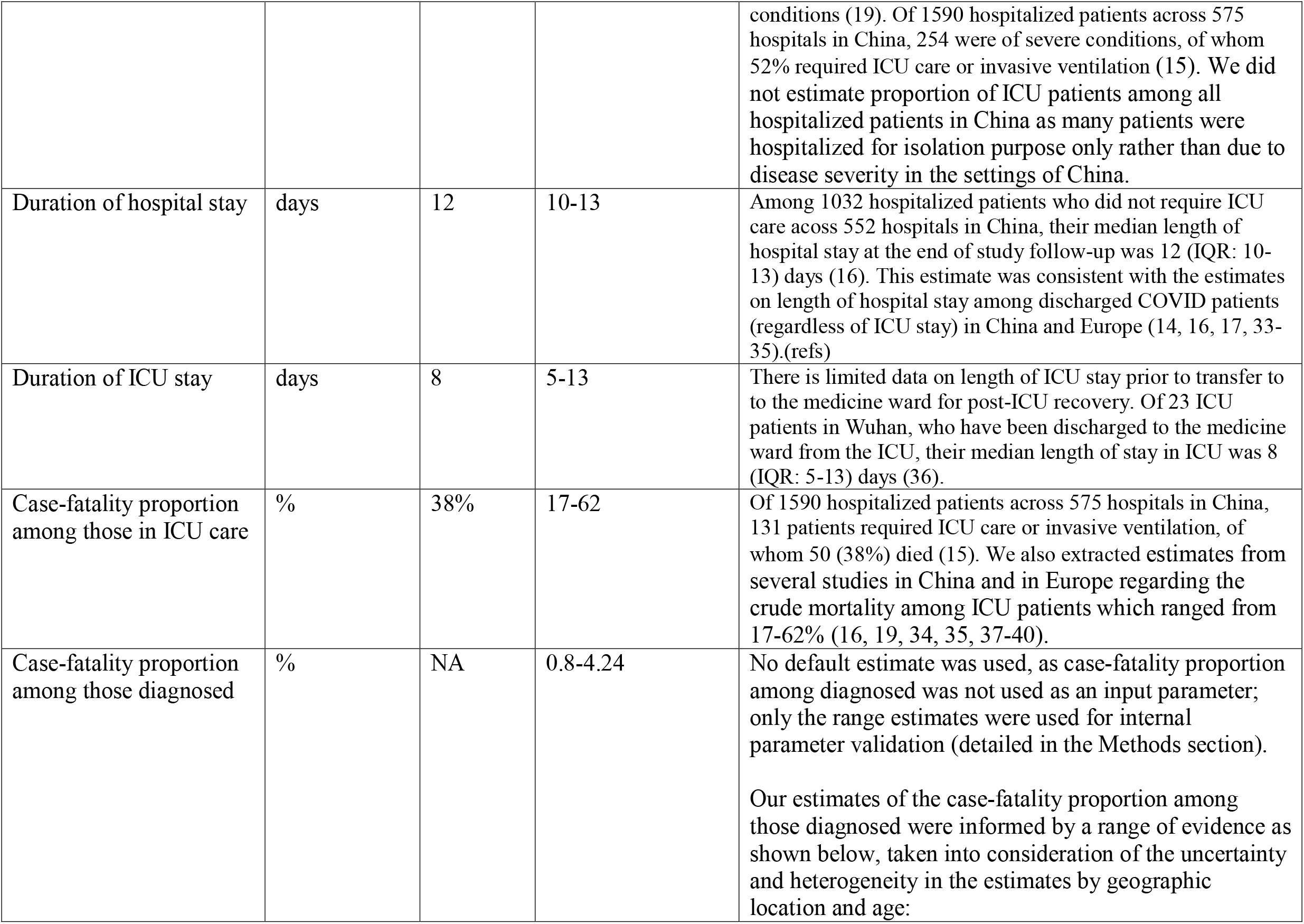
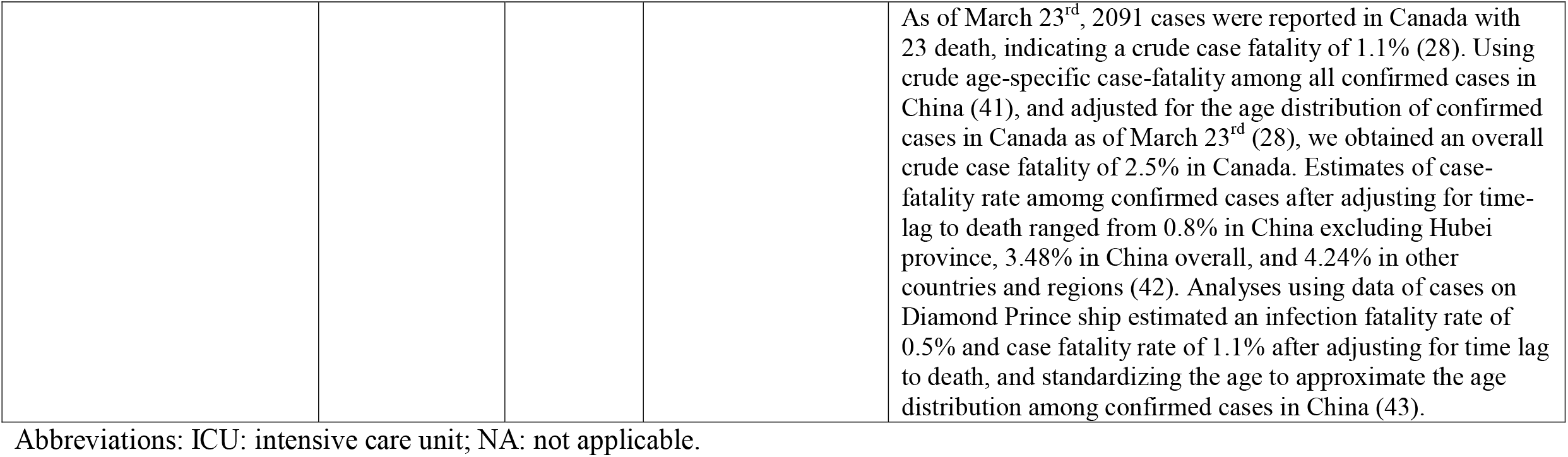
Transmission model parameters.

### *Hospital-specific estimates* (Appendix 1)

We used Institute for Clinical Evaluative Sciences (ICES) estimates on the median number (and inter-quartile range, IQR) of hospital admissions and intensive care unit (ICU) admissions in the GTA and at each hospital from March 2019 to August 2019 (12). Unity Health Toronto Decision Support provided daily census of non-ICU inpatients and ICU inpatients as a median (IQR) calculated over 90 days using March to June from the years 2014 to 2019 inclusive.

### Intervention parameters

We applied two interventions with assumptions surrounding their values: physical distancing to reduce contacts by 20% started 30 days into the outbreak; and the proportion of non-severe cases who self-isolate (default 10%). Intervention parameters were fixed for the primary analyses, and varied in sensitivity analyses (0 to 70% reduction in contact rate; delay initiating physical distancing from 2 to 90 days after start of outbreak; increasing the proportion with non-severe infection who self-isolate [following testing or syndromic diagnosis] from 10% to a the maximum proportion of individuals with COVID-19 who may develop symptoms (41-69% (21-23)).

### Epidemic constraints

To generate a plausible range of epidemic trajectories under best and worst-case scenarios, we sampled parameters as per **Table 1** while fixing the intervention parameters, and used the following constraints: the upper and lower bound of the per-capita, cumulative cases detected per day in Lombardy, Italy (24), and Hong Kong, China (25), respectively, within the first 30 days after detection of 3 cases. We then selected a slow/small epidemic and a fast/large epidemic using the lower and upper interquartile range in the peak incidence across the full, constrained set of epidemic trajectories. We defined a default scenario using the median or best-justified parameter values which passed our internal validity checks and epidemic constraints. We also examined the face validity of our default epidemic by comparing it to our synthesis the observed data in the GTA ((13-18, 26), **Appendix** 1).

### Analyses

First, we reported epidemic features and health care needs estimated by the range of plausible scenarios and the three selected scenarios for the GTA. Second, we applied GTA model outputs from the three scenarios to generate hospital-specific estimates using the catchment proportion for non-ICU and ICU hospital admissions and added the baseline daily (median) number of inpatients on all non-ICU and ICU units for each hospital. We then compared the potential trajectories, under the assumption that baseline admissions remain the same, with the maximum capacity for non-ICU and ICU beds at each hospital. Third, we performed a one-way sensitivity analysis using the default scenario to identify the main sources of uncertainty when estimating hospital surge.

### Ethics approval

This study was exempt from research ethics approval as the aggregate data provided by Unity Health Toronto Decision Support was not used to systematically investigate a hypothesis and thus, it was not considered human research as defined in TCPS2.

## RESULTS

**Figure 2** depicts the per-capita cumulative rate of confirmed cases across the plausible range of epidemics in the first 60 days of the outbreak, in the absence of further intervention. The default scenario follows a similar early trajectory of rapid growth in observed cases in the GTA, while the fast/large and slow/small epidemics are closer to, but not at the level of, Lombardy and Hong Kong, respectively (**Figure 2**).

**Figure 2.**
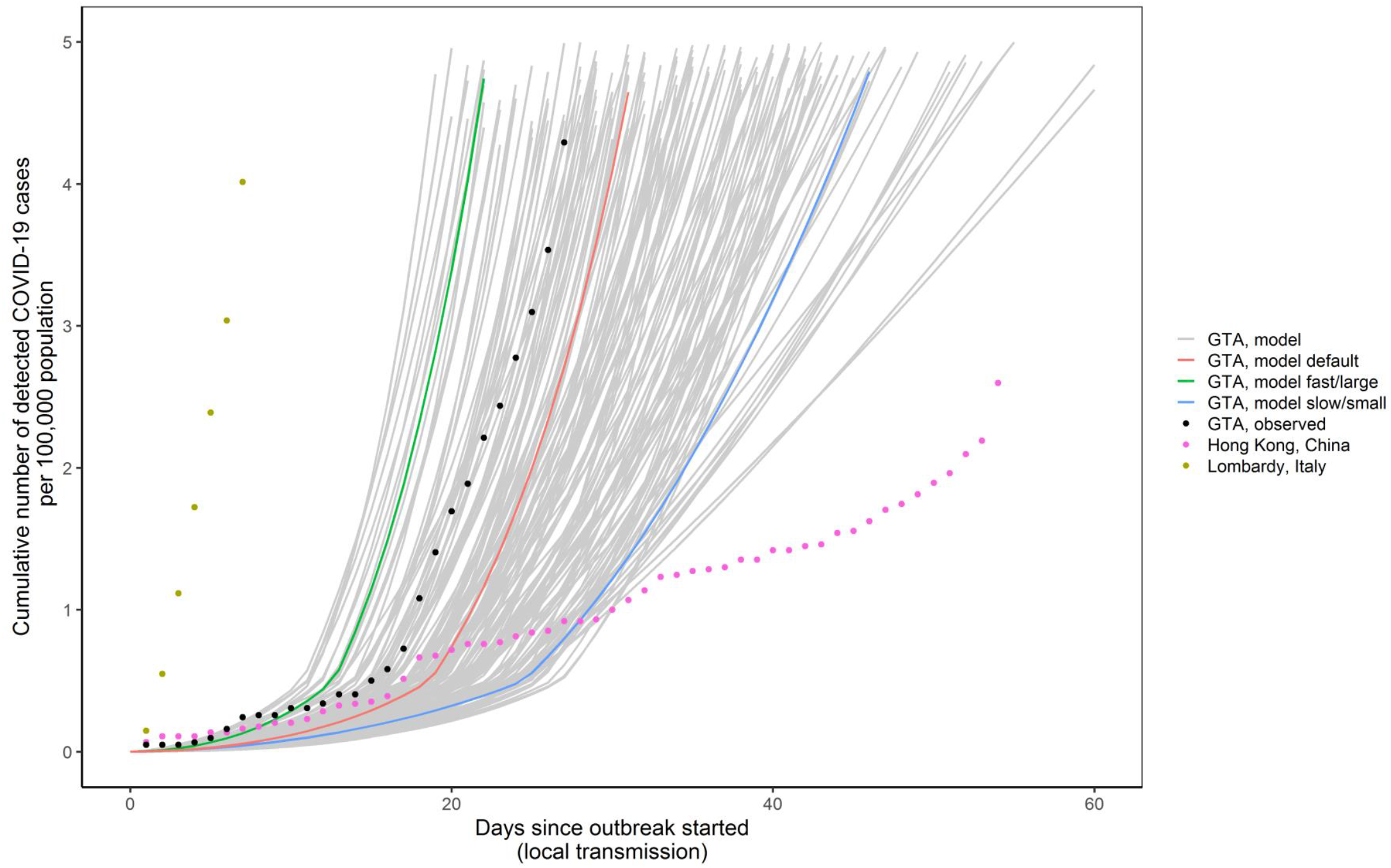
Cumulative detected cases per 100,000 population across simulated epidemic scenarios and observed data used for epidemic constraints. Model outputs from the sampled range of parameters in Table 1 which meet the model constraints are shown for detected cases as solid lines. The observed data for Lombardy, Italy and Hong Kong, China are shown as dotted lines, and the corresponding data points at day 30 since outbreak started were used as upper and lower bounds, respectively to constrain the epidemics. The observed data on cumulative detected cases for the Greater Toronto Area (travel-related, and local transmission) up to March 20, 2020 are also shown (dashed black line) as part of the face validity check. The model output for the fast/large epidemic is shown in green and slow/small epidemic in blue, selected as the upper and lower quartile of peak incidence, respectively, within the first 300 days. The default (solid red line) depicts the default scenario (Table 1). Simulated timeline begins at the start of the ‘seeding’ of the population with 0.0011-0.0048% of the population already infected with SARS-Cov-2. For observed data, we define outbreak started when 3 confirmed cases were observed. We chose 3 cases detected as the onset of epidemic based on the observed epidemic curve in the Greater Toronto Area, where the curve started to take off after detection of 3 cases. We applied the same threshold for other regions for comparability of epidemic curves across geographic locations. Abbreviations: GTA: Greater Toronto Area.

Parameter values for the three scenarios are compared in **Appendix Table 2.1**. The slow/small epidemic had a smaller R0: 1.84 vs. 2.4 in the default scenario. Transmission-related parameters were similar in the fast/large and default scenarios, except for a slightly higher proportion of the population already infected with COVID-19 at the start of the outbreak (initial seeding, 0.004% vs. 0.003% in the default scenario). However, cumulative confirmed cases (**Figure 2, Appendix 2 Figure 2**.**1**) were much lower in the default scenario because of the clinical parameters: the proportion of individuals with COVID-19 with severe disease requiring hospitalization, and thus, detected, was 10.4% in the fast/large vs. 5.5% in the default scenario.

**Table 2.**
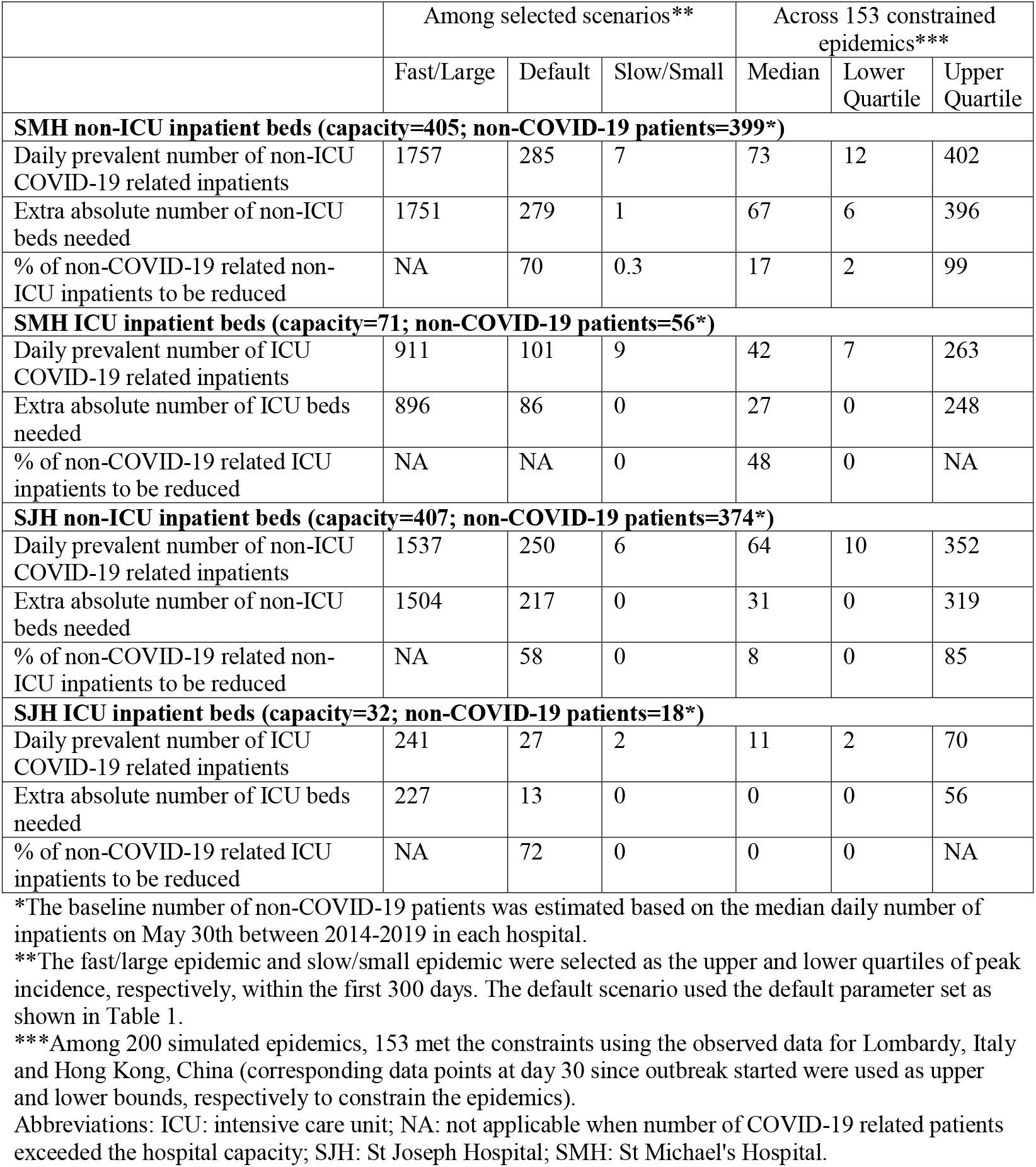
Prevalent number of baseline* inpatients and inpatients with COVID-19 in non-ICU and ICU beds in two acute care hospitals in the GTA, 90 days after outbreak started.

|  | Among selected scenarios** |  |  | Across 153 constrained epidemics*** |  |  |
| --- | --- | --- | --- | --- | --- | --- |
|  | Fast/Large | Default | Slow/Small | Median | Lower Quartile | Upper Quartile |
| **SMH non-ICU inpatient beds (capacity=405; non-COVID-19 patients=399\*)** |  |  |  |  |  |  |
| Daily prevalent number of non-ICU COVID-19 related inpatients | 1757 | 285 | 7 | 73 | 12 | 402 |
| Extra absolute number of non-ICU beds needed | 1751 | 279 | 1 | 67 | 6 | 396 |
| % of non-COVID-19 related non-ICU inpatients to be reduced | NA | 70 | 0.3 | 17 | 2 | 99 |
| **SMH ICU inpatient beds (capacity=71; non-COVID-19 patients=56\*)** |  |  |  |  |  |  |
| Daily prevalent number of ICU COVID-19 related inpatients | 911 | 101 | 9 | 42 | 7 | 263 |
| Extra absolute number of ICU beds needed | 896 | 86 | 0 | 27 | 0 | 248 |
| % of non-COVID-19 related ICU inpatients to be reduced | NA | NA | 0 | 48 | 0 | NA |
| **SJH non-ICU inpatient beds (capacity=407; non-COVID-19 patients=374\*)** |  |  |  |  |  |  |
| Daily prevalent number of non-ICU COVID-19 related inpatients | 1537 | 250 | 6 | 64 | 10 | 352 |
| Extra absolute number of non-ICU beds needed | 1504 | 217 | 0 | 31 | 0 | 319 |
| % of non-COVID-19 related non-ICU inpatients to be reduced | NA | 58 | 0 | 8 | 0 | 85 |
| **SJH ICU inpatient beds (capacity=32; non-COVID-19 patients=18\*)** |  |  |  |  |  |  |
| Daily prevalent number of ICU COVID-19 related inpatients | 241 | 27 | 2 | 11 | 2 | 70 |
| Extra absolute number of ICU beds needed | 227 | 13 | 0 | 0 | 0 | 56 |
| % of non-COVID-19 related ICU inpatients to be reduced | NA | 72 | 0 | 0 | 0 | NA |

\*The baseline number of non-COVID-19 patients was estimated based on the median daily number of inpatients on May 30th between 2014-2019 in each hospital.
\*\*The fast/large epidemic and slow/small epidemic were selected as the upper and lower quartiles of peak incidence, respectively, within the first 300 days. The default scenario used the default parameter set as shown in Table 1.
\*\*\*Among 200 simulated epidemics, 153 met the constraints using the observed data for Lombardy, Italy and Hong Kong, China (corresponding data points at day 30 since outbreak started were used as upper and lower bounds, respectively to constrain the epidemics).
Abbreviations: ICU: intensive care unit; NA: not applicable when number of COVID-19 related patients exceeded the hospital capacity; SJH: St Joseph Hospital; SMH: St Michael's Hospital.

### Comparing scenarios for hospital surge within the GTA

**Figure 3a** shows the epidemic curves in the absence of further interventions. Given the similar transmission parameters, the default and fast/large epidemic follow similar underlying patterns. As such, the default scenario represents a pessimistic scenario with 71.0% of the population infected by day 300.

**Figure 3.**
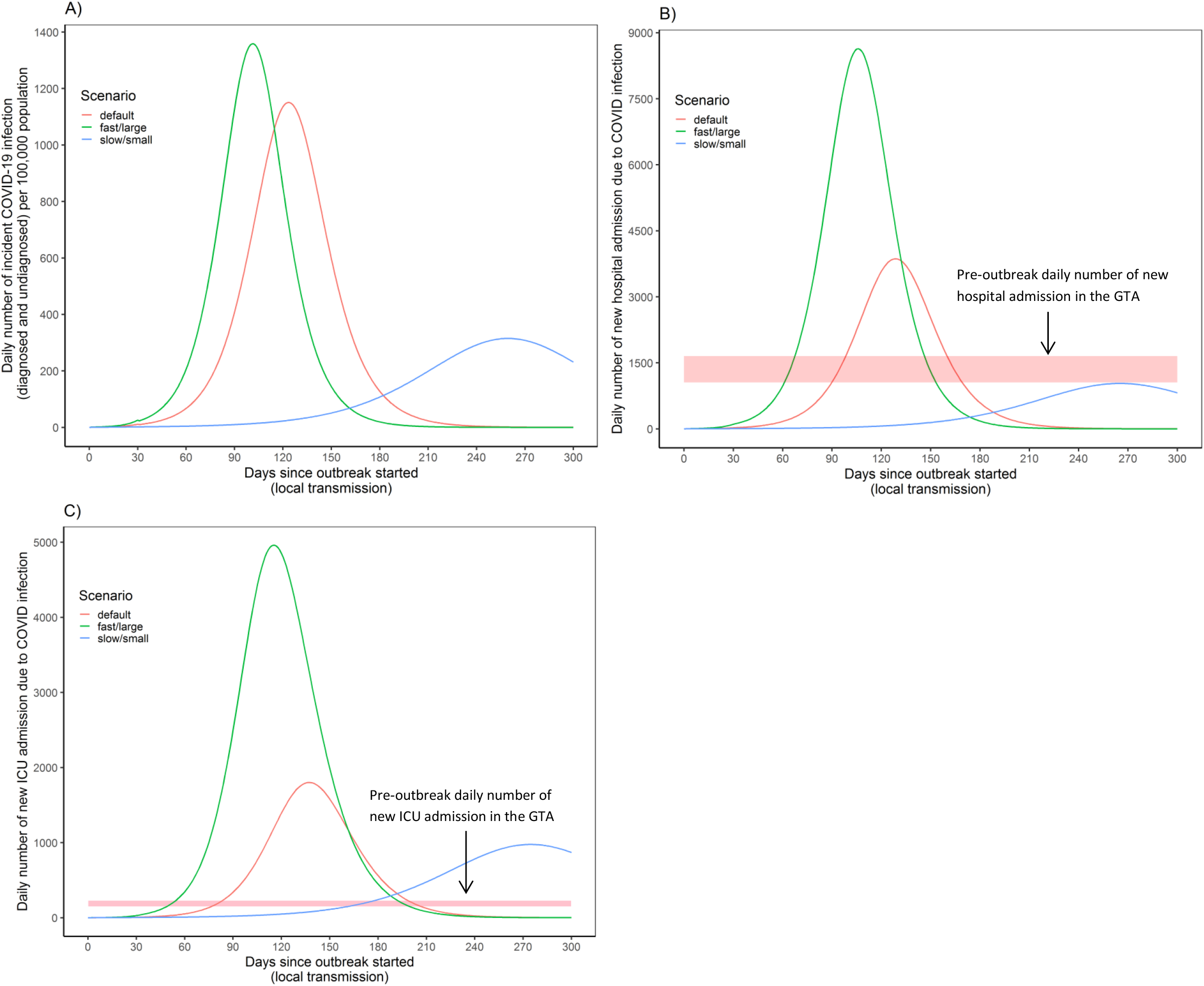
Incident epidemic curves and health-care needs in the Greater Toronto Area across three scenarios: default, fast/large, slow/small epidemics. (A) modeled incidence of infection (diagnosed and undiagnosed) for the GTA; (B) modeled daily number of hospital admissions for individuals with COVID-19 alongside pre-outbreak data on the daily median number of hospital admissions between March-August 2019 in the GTA; (C) modeled daily number of ICU admissions alongside pre-outbreak data on the daily median number of ICU admissions between March-August 2019 in the GTA. The fast/large and slow/small epidemic scenarios are selected based on the upper and lower quartile of peak incidence, respectively, within the first 300 days across all simulated constrained epidemics; the default scenario reflects default parameter set in Table 1. All three scenarios (default, fast/large, and slow/small) assume that physical distancing started on day 30 and reduced contact rates by 20%, but has not increased nor decreased thereafter; and that the proportion of individuals with non-severe COVID-19 who self-isolate (e.g. via diagnosis of confirmed/suspected COVID-19) has not changed over the course of the epidemic. Abbreviations: GTA: greater Toronto area; ICU: intensive care unit.

In 2019, an estimated 1,056 to 1,653, and 145 to 231 patients were admitted each day to a non-ICU bed (**Figure 3b**) and to an ICU bed (**Figure 3c**) respectively. Despite similar underlying epidemics, the fast/large and default epidemics project different health-care needs driven by differences in probability of severe disease. In the absence of further interventions, ICU admissions in the small/slow epidemic still surpass the daily number of ICU admissions in 2019 (**Figure 3c**).

**Appendix 2 Table 2**.**2** summarizes the peak number of admissions, and peak in daily census (prevalence) of inpatients within the first 300 days of the outbreak. Across all plausible scenarios, the IQR of peak prevalence in number of non-ICU inpatients with COVID-19 ranges between 10,189 and 38,502; and for ICU inpatients ranges between 2,454 and 17,651. In the default scenario, the model estimates a peak of 32,368 non-ICU and 7,418 ICU inpatient beds needed to care for patients with COVID-19 in the GTA.

### Hospital-specific surge

Between March to August 2019, St. Michael’s Hospital and St. Joseph’s Hospital, respectively, received 4.5% (95% CI 4.4, 4.6) and 3.9% (95% CI 3.8, 4.0) of all non-ICU hospital admissions in the GTA; and 8.7% (95% CI 8.4, 9.0) and 2.3% (95% CI 2.1-2.5) of ICU admissions in the GTA. In the years from 2014-2019, the median daily non-ICU and ICU inpatient census at St. Michael’s Hospital was 370-419 and 50-59, with a maximum capacity of 405 and 71 beds, respectively (**Appendix 1**). At St. Joseph’s Health Centre, the median daily non-ICU and ICU inpatient census was 353-390 and 17-23, with a maximum capacity of 407 and 32 beds, respectively (**Appendix 1**).

Thus, the total daily census of non-ICU and ICU inpatients, with or without COVID-19 is shown in **Figure 4 and Figure 5** respectively for St. Michael’s Hospital, and in **Appendix 2** for St. Joseph’s Health Centre (**Figure 2**.**2 and 2**.**3**). The model estimates that if nothing changes with the baseline (pre-outbreak) levels of admissions, both hospitals will surpass non-ICU and ICU capacity under the fast/large and default scenarios within 90 days of the outbreak, but (as expected based on **Figure 3c**) that may not be the case with the small/slow epidemic (**Figures 4-5, and Appendix 2 Figure 2**.**2-2**.**3**). Driven by differences in their catchment, St. Michael’s Hospital may expect an earlier surge around day 40 and St. Joseph’s Health Centre, a later surge around day 65.

**Figure 4.**
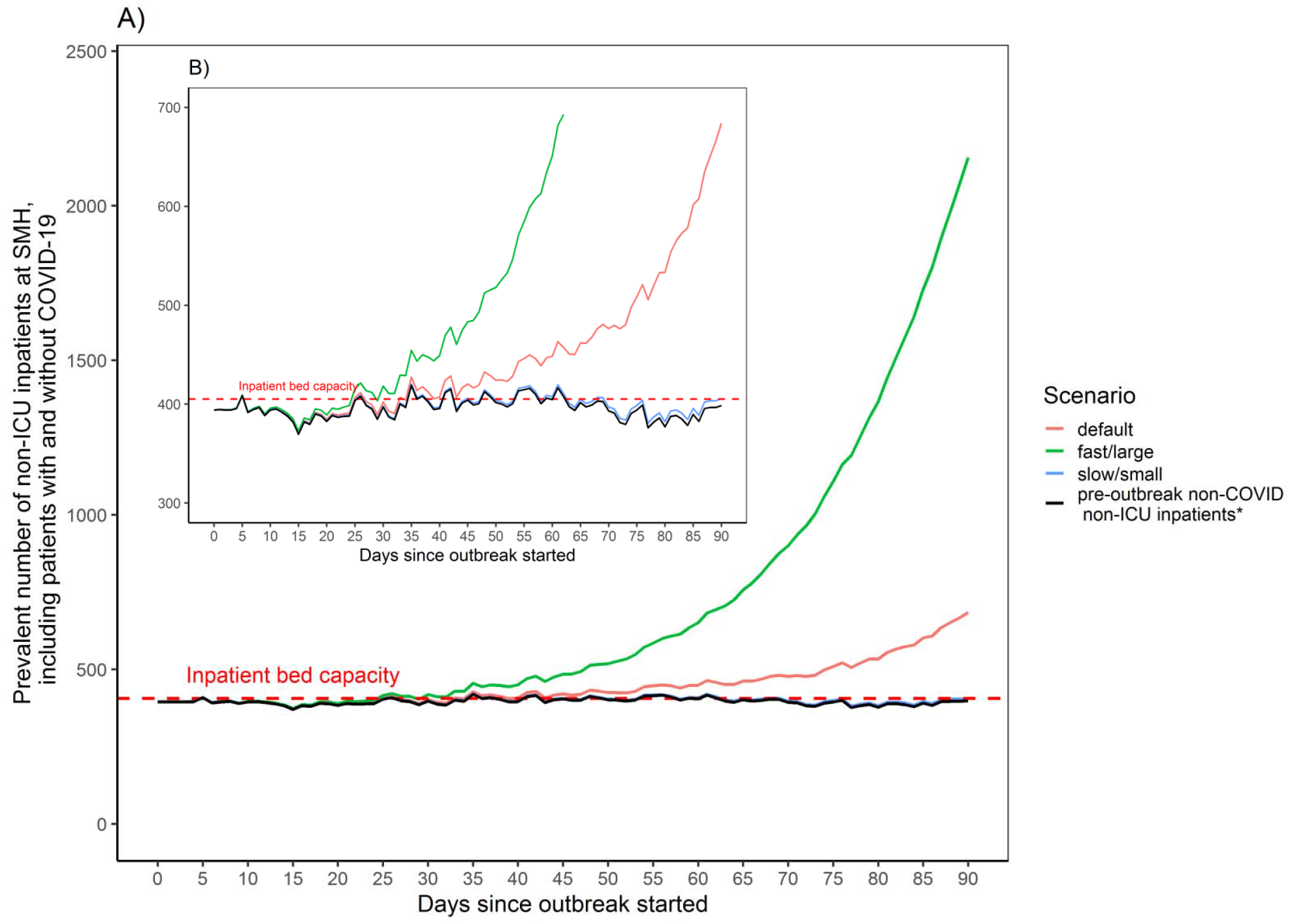
Estimated surge and capacity for non-ICU hospitalization at St. Michael’s Hospital in the Greater Toronto Area. (A) Modeled number of non-ICU inpatients (including inpatients with and without COVID-19) and corresponding pre-outbreak baseline (non-COVID) number of non-ICU inpatients per day over 90 days. *Estimated by the median number of non-ICU inpatients at SMH between March – June, 2014-2019. (B) Same information as (A) but the y-axis ranged between 300-700. Estimates assume that distribution of non-ICU hospital admissions for patients with COVID-19 follows the pre-outbreak catchment of all non-ICU admissions across acute care hospitals in the GTA (March – August 2019), such that St. Michael’s Hospital receives 4.5% of all non-ICU hospital admissions. Our use of observed data on hospital-specific non-ICU admissions during March-June (black line) are not meant to indicate a start-date of the outbreak as March 1. All three scenarios (default, fast/large, and slow/small) assume that physical distancing started on day 30 and reduced contact rates by 20%, but has not increased nor decreased; and that the proportion of individuals with non-severe COVID-19 who self-isolate (e.g. via diagnosis of confirmed/suspected COVID-19) has not changed over the course of the epidemic. Abbreviations: ICU: intensive care unit; SMH: St. Michael’s Hospital.

**Figure 5.**
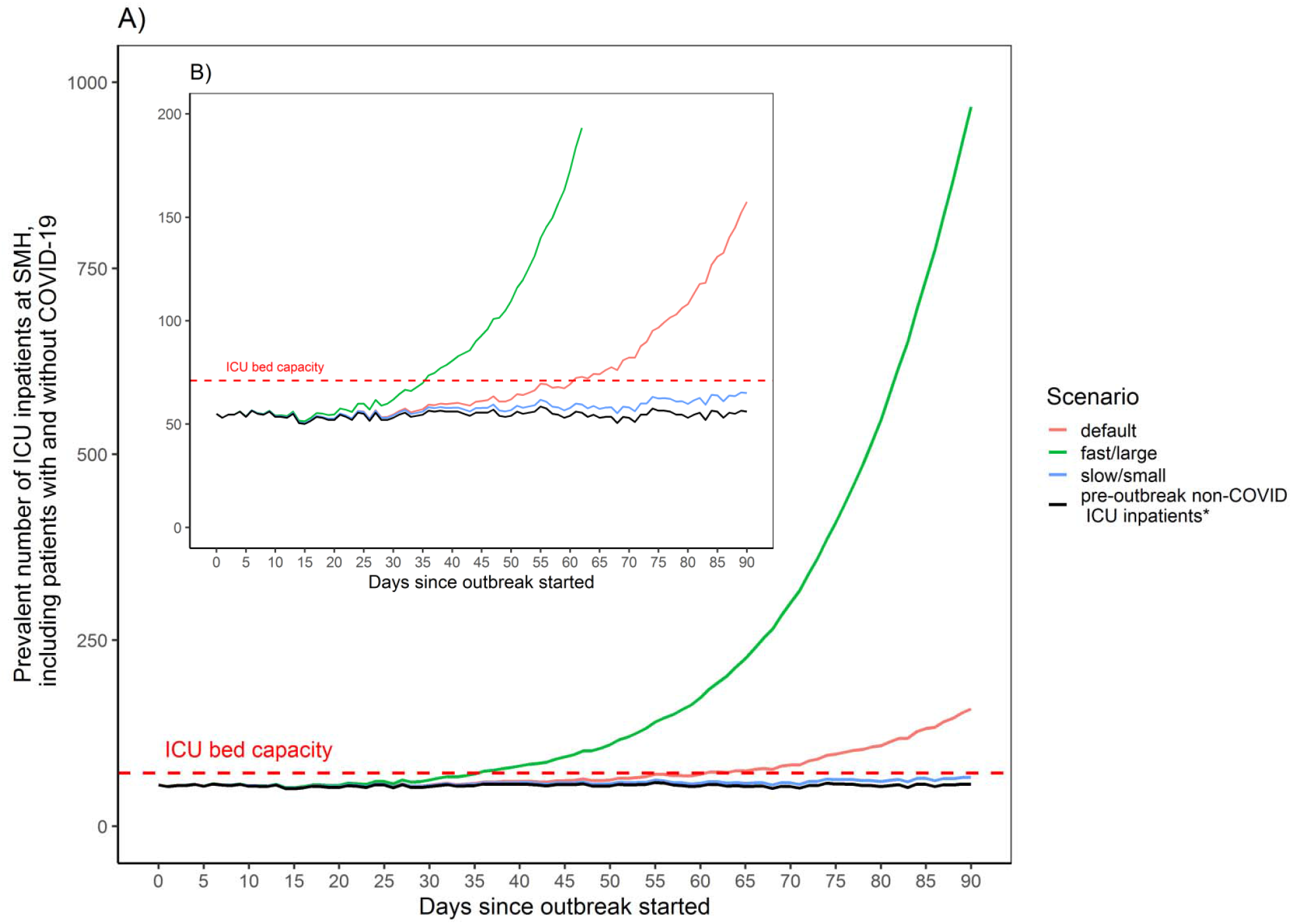
Estimated surge and capacity for ICU care at St. Michael’s Hospital in the Greater Toronto Area. (A) Modeled number of ICU inpatients (including inpatients with and without COVID-19) and corresponding pre-outbreak baseline (non-COVID) number of ICU inpatients. *Estimated by the median number of ICU inpatients at SMH between March – June, 2014-2019). (B) Same information as (A) but the y-axis ranged between 300-700. Estimates assume that distribution ICU admissions for patients with COVID-19 follows the pre-outbreak catchment of all ICU admissions across acute care hospitals in the GTA (March – August 2019), such that St. Michael’s Hospital receives 8.7% of all ICU hospital admissions. Our use of observed data on hospital-specific ICU admissions during March-June (black line) are not meant to indicate a start-date of the outbreak as March 1. All three scenarios (default, fast/large, and slow/small) assume that physical distancing started on day 30 and reduced contact rates by 20%, but has not increased nor decreased; and that the proportion of individuals with non-severe COVID-19 who self-isolate (e.g. via diagnosis of confirmed/suspected COVID-19 or without) has not changed over the course of the epidemic. Abbreviations: ICU: intensive care unit; SMH: St. Michael’s Hospital.

**Table 2** provides the daily census (prevalence) of inpatients with COVID-19 from each scenario, the median and IQR of the full range of constrained model outputs for the catchment of each hospital, and the relative reduction in non-COVID admissions or absolute increase in ICU beds needed to address the surge at each site. In the default scenario, for St. Michael’s Hospital to remain below its non-ICU bed capacity 90 days into the outbreaks, the hospital would need to reduce non-ICU inpatient care for non-COVID by 70%, to open up 279 non-ICU inpatient beds; St. Michael’s Hospital would also need to create 86 new ICU-beds in addition to its current capacity of 71 beds to be able to care for non-COVID and new COVID-related ICU inpatients (**Table 2**). At St. Joseph’s Health Centre, under the default scenario, non-ICU beds and ICU-beds for non-COVID would need to be reduced by 58% and 72%, respectively, to open up 217 non-ICU beds and 13 ICU beds by 90 days into the outbreak, to remain below the hospital’s respective bed capacity (**Table 2**).

### Sensitivity analyses

Results of one-way sensitivity analyses using the default scenario, for non-ICU and ICU care are shown for St. Michael’s Hospital in **Figure 6** and **Appendix 2 Figures 2**.**4-2**.**7**, respectively. Results of sensitivity analyses were similar for St. Joseph’s Health Centre. At the hospital-level, uncertainty in local epidemiological features was more influential than uncertainty in clinical severity. For example, uncertainty in local seeding **(Figure 6a)** has a larger influence on non-ICU care at St. Michael’s Hospital than uncertainty in clinical severity (**Figure 6b**). The effect of early versus delayed initiation of physical distancing has a large impact as shown in **Figure 6c**. If physical distancing could only reduce contact rates by 20%, then maximizing the diagnostic capacity or syndromic diagnosis at the community-level in the GTA could reduce the surge at St. Michael’s hospital from 285 to 40 non-ICU patients with COVID-19 and 101 to 10 ICU patients with COVID-19 by day 90 of the outbreak.

**Figure 6.**
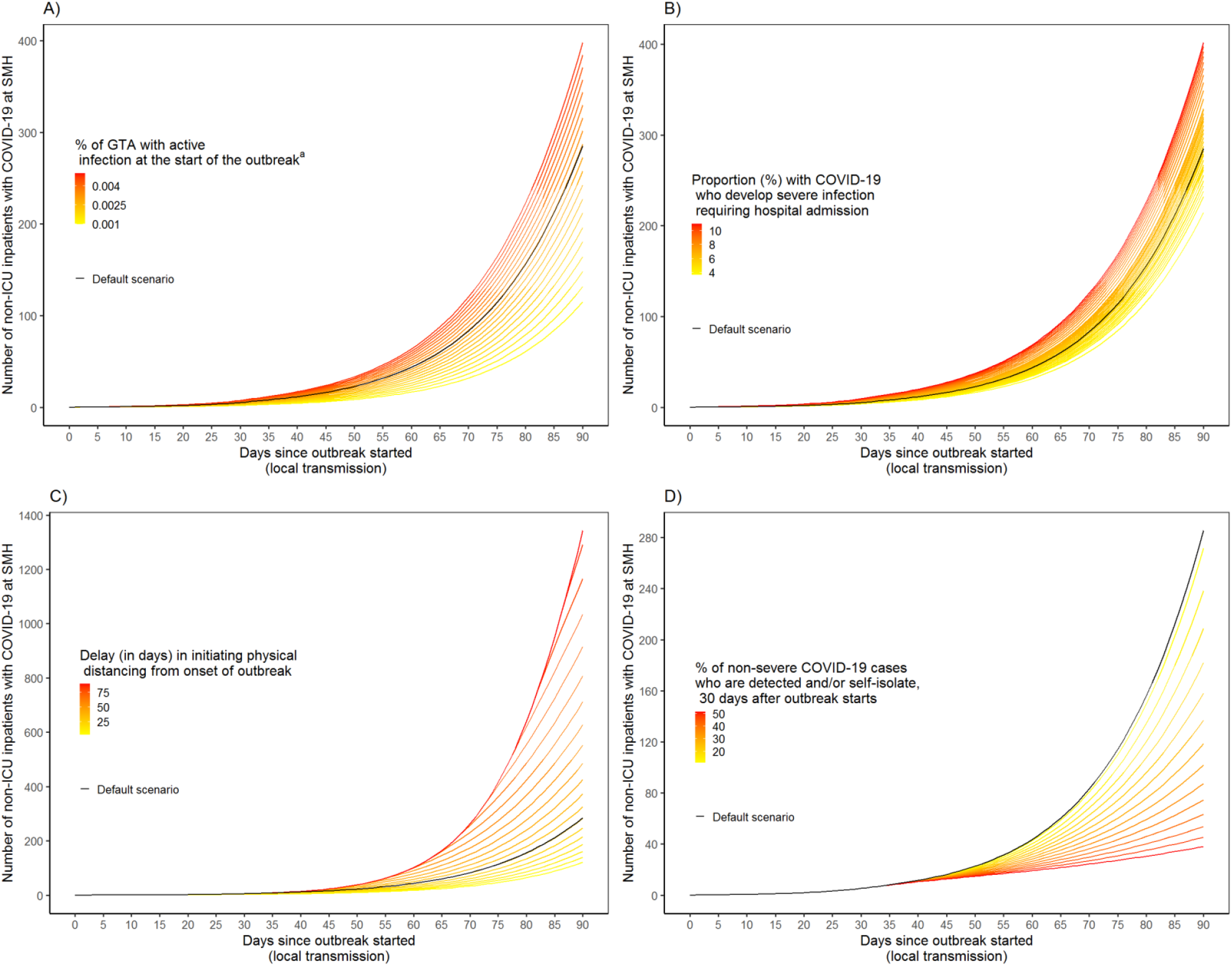
One-way sensitivity analyses using default epidemic scenario for prevalence of non-ICU inpatients with COVID-19 at St. Michael’s Hospital. The influence of (A) seeding (proportion of population already infected with COVID-19 just at the start of the outbreak); and (B) clinical severity (proportion of individuals infected with COVID-19 who require hospitalization); (C) earlier or later initiation of physical distancing (from start of outbreak to 60 days after outbreak started); and (D) proportion of individuals with non-severe COVID-19 who are diagnosed and/or self-isolate, 30 days after outbreak starts (e.g. due to increase capacity in testing in the community). Note that the y-axis scales for figures are different. Abbreviations: ICU: intensive care unit; SMH: St Michael’s Hospital; GTA: Greater Toronto Area.

## DISCUSSION

In the absence of further interventions, even a best-case scenario like the simulated small/slow epidemic, may lead to a surge in ICU care in the city. However, the impact of the city’s outbreak is expected to vary across hospitals by their local catchment, with local epidemic features driving each hospital’s surge. The local transmission dynamics, or what was happening with the epidemic overall in the city, had a larger influence on a hospital’s surge than uncertainty around disease severity. As such, community-level interventions, like maximizing diagnosis (via testing, or via syndromic case finding) among symptomatic individuals in the community could potentially mitigate the surge in each hospital.

Our estimates of the surge at the hospital-level align with the relative magnitude of surge at a macro-level as estimated from other modeling studies (provincial and national (27) in Canada, and in other settings (28)), but add to the literature by demonstrating potential variability with even minimal variability in hospital-context. The preliminary hospital-specific findings (on March 4, 2020) were used to prepare for the local surge at the two hospitals. First, the hospitals opened up beds by temporarily cancelling non-essential surgeries and procedures. Second, as most COVID-related inpatient care would fall under the hospitalist and medicine services, the relevant departments rapidly set up a separate service with a viable back-up system and ability for rapid scale up in anticipation of increasing cases requiring admission, and staffing short-falls due to infection, exposure, or while awaiting test results if symptomatic. Third, ambulatory clinics were reduced with a focus on virtual care and urgent assessments only; this allowed clinic space to be consolidated to preserve personal protective equipment and human resources (including physicians) for deployment to other areas. This consolidation also allowed identification of potential inpatient spaces. There was also a change in health-care use by the public: non-COVID medicine admissions are dropping across the city and country (29). Thus, the next iteration of analyses will need to account for active and passive reductions in admissions.

Limitations include our assumption that the distribution of hospitalizations and ICU admissions would follow 2019 patterns, and that transmission was homogenous across the city. However, distribution of admissions may be expected to follow even more granular patterns of transmission in the hospital’s neighborhood-level catchment area (30). Future work includes capturing heterogeneity within the five health units and near real-time adjustment of the catchment using observed patterns of hospital-specific admissions. Finally, our objective was to conduct a scenario-based analyses, and not to explicitly fit the model to observed cases, hospitalizations, ICU admissions and deaths in the GTA; these are the next step in supporting local GTA hospitals and re-distribution of ICU care across the city (31).

In summary, a surge in hospital capacity in the GTA is expected across a range of pessimistic to optimistic scenarios during the COVID-19 pandemic, with important and practical variability anticipated at the hospital-level. What is happening outside the hospital will have the largest influence on each hospital’s surge, with an opportunity for increasing diagnostic (testing or syndromic) capacity to mitigate each hospital’s surge, especially if there are pragmatic constraints on physical distancing measures. ICU admissions at the city-level is expected to surge past baseline even in best-case scenarios, but with variability across hospitals – thus, signaling the importance of efforts to plan and redistribute ICU care with where variability in surge may be expected.

## Data Availability

Model codes and data are available at: https://github.com/mishra-lab/covid-GTA-surge-planning.

## DATA SHARING

Model codes and data are available at: https://github.com/mishra-lab/covid-GTA-surge-planning.

## ACKNOWLEDGMENTS

We thank the Infection Prevention and Control practitioners and Pandemic Command Centers at every health-facility across the GTA who have come together as a working group, and whose efforts, solidarity, and tireless work within each of their hospitals, guided the questions in this study. In particular, we thank the Infection Prevention and Control Teams and Pandemic Command Centre at Unity Health Toronto for requesting and guiding the questions.

We thank Jesse Knight for helpful feedback on the R Shiny tool.

This study was supported by ICES, a non-profit research institute funded by the Ontario Ministry of Health. Parts of this material are based on data and information compiled and provided by the Ontario Ministry of Health and the Canadian Institute for Health Information. The analyses, conclusions, opinions and statements expressed herein are those of the authors and not necessarily those of the funding or data sources; no endorsement is intended or should be inferred.

The study was also supported by Unity Health Toronto Decision Support Analyses. We thank Pavidra Ambiganithy, St. Michael’s Hospital Infection Prevention and Control Team, for supporting the coordination surrounding data requests.

The transmission modeling study was supported by the Canadian Institutes of Health Research Foundation Grant FN 13455, and the Ontario Early Researcher Award Number ER17-13-043.

SM is supported by Tier 2 Canada Research Chair in Mathematical Modeling and Program Science.

SS is supported by a Tier 1 Research Chair in Knowledge Translation and Quality of Care.

